# Association of upper respiratory *Streptococcus pneumoniae* colonization with SARS-CoV-2 infection among adults

**DOI:** 10.1101/2022.10.04.22280709

**Authors:** Anna M. Parker, Nicole Jackson, Shevya Awasthi, Hanna Kim, Tess Alwan, Anne L. Wyllie, Alisha B. Baldwin, Nicole B. Brennick, Erica A. Moehle, Petros Giannikopoulos, Katherine Kogut, Nina Holland, Ana Mora-Wyrobek, Brenda Eskenazi, Lee W. Riley, Joseph A. Lewnard

## Abstract

**Background:** *Streptococcus pneumoniae* interacts with numerous viral respiratory pathogens in the upper airway. It is unclear whether similar interactions occur with SARS-CoV-2.

**Methods:** We collected saliva specimens from working-age adults receiving SARS-CoV-2 molecular testing at outpatient clinics and via mobile community-outreach testing between July and November 2020 in Monterey County, California. Following bacterial culture enrichment, we tested for pneumococci by quantitative polymerase chain reaction (qPCR) targeting the *lytA* and *piaB* genes, and measured associations with SARS-CoV-2 infection via conditional logistic regression.

**Results:** Analyses included 1,278 participants, with 564 enrolled in clinics and 714 enrolled through outreach-based testing. Prevalence of pneumococcal carriage was 9.2% (117/1,278) among all participants (11.2% [63/564] clinic-based testing; 7.6% [54/714] outreach testing). Prevalence of SARS-CoV-2 infection was 27.4% (32/117) among pneumococcal carriers and 9.6% (112/1,161) among non-carriers (adjusted odds ratio [aOR]: 2.73; 95% confidence interval: 1.58-4.69). Associations between SARS-CoV-2 infection and pneumococcal carriage were enhanced in the clinic-based sample (aOR=4.01 [2.08-7.75]) and among symptomatic participants (aOR=3.38 [1.35-8.40]), when compared to findings within the outreach-based sample and among asymptomatic participants. Adjusted odds of SARS-CoV-2 co-infection increased 1.24 (1.00-1.55)-fold for each 1-unit decrease in *piaB* qPCR C_*T*_ value among pneumococcal carriers. Last, pneumococcal carriage modified the association of SARS-CoV-2 infection with recent exposure to a suspected COVID-19 case (aOR=7.64 [1.91-30.7] and 3.29 [1.94-5.59]) among pneumococcal carriers and non-carriers, respectively).

**Conclusions:** Associations of pneumococcal carriage detection and density with SARS-CoV-2 suggest a synergistic relationship in the upper airway. Longitudinal studies are needed to determine interaction mechanisms between pneumococci and SARS-CoV-2.

**Key points:**

- In an adult ambulatory and community sample, SARS-CoV-2 infection was more prevalent among pneumococcal carriers than non-carriers.
- Associations between pneumococcal carriage and SARS-CoV-2 infection were strongest among adults reporting acute symptoms and receiving SARS-CoV-2 testing in a clinical setting.

## INTRODUCTION

Interactions involving *Streptococcus pneumoniae* and respiratory viruses feature in the natural history of multiple clinical conditions. Pneumococcal pneumonia and invasive pneumococcal disease (IPD) incidence rates align with seasonal influenza and respiratory syncytial virus (RSV) transmission,^1,2^ and severe pneumococcal infections are prominent sequelae of viral respiratory disease.^3^ Interactions also arise earlier in the clinical course, including when symptoms are absent. Epidemiologic studies of influenza virus, RSV, human rhinoviruses, and endemic human coronaviruses have revealed frequent co-detection of pneumococci in the upper airway, often in association with increased density of bacterial shedding.^4–11^ Likewise, experimental challenge models in animals^12^ and humans^13,14^ have revealed effects of pneumococcal carriage on influenza virus acquisition and immune response, while epidemiologic studies have identified increased risk of viral acquisition and acute respiratory infection associated with the presence and density of pneumococcal carriage.^15^

It is unclear whether similar interactions occur between pneumococci and SARS-CoV-2. Contrary to experience with influenza, early investigations identified low risk of severe secondary pneumococcal infections among COVID-19 patients,^16^ although such studies may have been limited by choice of pneumococcal detection methods and prevalent antibiotic prescribing for COVID-19 patients during the early pandemic.^17,18^ Among healthcare workers in the United Kingdom, SARS-CoV-2 infection was associated with three-fold higher likelihood of molecular detection of pneumococcal carriage in saliva; moreover, pneumococcal carriers experienced diminished IgA and CD4^+^ T-cell responses to SARS-CoV-2 after recovery.^19^ SARS-CoV-2 acquisition concurrent with or shortly following IPD was associated with enhanced mortality in a UK surveillance study.^20^ Last, among adults aged ≥65 years in the United States, receipt of 13-valent pneumococcal conjugate vaccine (PCV13) was associated with reduced risk of COVID-19 diagnosis, hospital admission, and mortality.^21^ These findings reflect moderate PCV effectiveness against virus-associated lower respiratory tract infection (LRTI) and pneumonia among children and adults in prior studies.^22–25^

To understand potential interactions between pneumococci and SARS-CoV-2, we evaluated pneumococcal carriage within a study of SARS-CoV-2 infection among adult farmworkers in Monterey County, California undertaken between July and November, 2020.^26^ We aimed to assess whether pneumococcal carriage was associated with SARS-CoV-2 infection, and to evaluate how this association varied according to pneumococcal carriage density and clinical symptoms.

## METHODS

### Study setting and recruitment

Primary study methods and findings have been reported previously.^26–28^ Briefly, we enrolled adults currently or previously employed in agricultural work through no-cost, point-of-care molecular SARS-CoV-2 testing at ambulatory outpatient clinics and mobile community outreach venues operated by Clínica de Salud del Valle de Salinas (CSVS) throughout Monterey County, California, from 16 July to 30 November, 2020. CSVS is a Federally-Qualified Health Center serving >50,000 patients, primarily focused on healthcare for local agricultural workers and their families. Most farmworkers in the region are Latino and live below the federal poverty line.^29^ SARS-CoV-2 test positivity among these farmworkers was higher than among other adults in Monterey County throughout the study period.^26^

Healthcare providers at CSVS recommended SARS-CoV-2 testing for all patients, regardless of symptoms, during the study period. Outreach testing was also conducted at community sites throughout the Salinas Valley, including low-income and agricultural employer-provided housing facilities, agricultural fields, and health fairs. Individuals eligible for the primary study were nonpregnant adult farmworkers aged ≥18 years who received SARS-CoV-2 testing at CSVS, who had not previously participated in the study, and who spoke sufficient Spanish or English to provide consent and complete study procedures. For this ancillary study of pneumococcal carriage, we expanded recruitment to include any other adults (not exclusively farmworkers) who received SARS-CoV-2 testing at CSVS, who spoke Spanish or English and agreed to provide a saliva sample.

Participants provided ≥3 mL of saliva passively pooling in the front of the mouth into ≥20 mL screw-top collection tubes. Participants responded to a standardized survey covering demographics, clinical symptoms, and recent exposures by telephone interview within 48 hours of their testing appointment, before SARS-CoV-2 testing results were notified. Participants who were not farmworkers and enrolled in the pneumococcal sub-study only answered a shortened survey.

### Outcome definitions

We determined pneumococcal carriage via quantitative polymerase chain reaction (qPCR) assays targeting the *lytA* and *piaB* genes, using bacterial DNA extracted after a culture-enrichment step according to previously-validated protocols^30^ (**Text S1**). Samples with *lytA* cycle threshold (C_*T*_) values <40 were tested for the *piaB* gene and were considered to contain pneumococci if both *lytA* and *piaB* C_*T*_ values were <40 (**Table S1**). We additionally report results applying a more stringent threshold of *lytA* and *piaB* C_*T*_ values <35. Clinical testing for SARS-CoV-2 from oropharyngeal specimens was conducted via the qualitative Aptima SARS-CoV-2 (Hologic; Marlborough, Massachusetts, US) transcription-mediated amplification (TMA) assay. We considered individuals with positive results for SARS-CoV-2 by this clinical test to be infected. For secondary analyses addressing SARS-CoV-2 viral abundance, we also used C_*T*_ values from qPCR-based detection of the SARS-CoV-2 nucleocapsid (N) and structural envelope protein (E) genes in saliva specimens (**Text S1**).

### Statistical analysis

We first compared prevalence of SARS-CoV-2 infection among pneumococcal carriers and non-carriers. We fit conditional logistic regression models defining SARS-CoV-2 infection as the outcome and pneumococcal carriage as the primary exposure of interest, with strata for participants’ recruitment setting (clinic-based testing or mobile outreach testing) to address differences in the populations reached. We computed adjusted odds ratios (aORs) adjusting for risk factors including participants’ age, sex, body mass index (BMI), community of residence, current or former smoking, residence type, educational background, household annual income, language spoken, exposure to children aged <5 years, and potential COVID-19 related exposures and risk behaviors in the prior two weeks. These included contact with a confirmed or suspected COVID-19 case, mask-wearing in indoor public settings, attendance at indoor gatherings with individuals from outside participants’ households, and regular handwashing. We repeated analyses within two sets of subgroups: participants recruited in clinic-based or mobile outreach testing venues, and participants who reported experiencing or not experiencing clinical symptoms within 2 weeks before testing. As secondary analyses, we also fit models defining pneumococcal carriage as *lytA* and *piaB* C_*T*_ <35.

To determine if associations varied according to pneumococcal density, we fit conditional logistic regression models partitioning pneumococcal carriage according to *lytA* and *piaB* C_*T*_ in the ranges of 35-39.9, 30-34.9, and <30. We also fit models subset to participants carried pneumococci, within which we defined *lytA* and *piaB* C_*T*_ as continuous quantitative covariates. To determine if SARS-CoV-2 abundance was associated with pneumococcal detection or density, we also fit regression models relating C_*T*_ values (as continuous outcomes) for the SARS-CoV-2 N and E genes in saliva to pneumococcal detection. Each of these analyses adjusted for the covariates included in primary analyses.

Last, we sought to determine whether the association between recent SARS-CoV-2 exposure and risk of infection differed in association with pneumococcal carriage detection. We conducted analyses considering two distinct SARS-CoV-2 exposure definitions: self-reported contact with any individual experiencing acute upper respiratory symptoms (“suspected” exposure; expected to provide a sensitive albeit non-specific indicator), and notification of exposure to a confirmed SARS-CoV-2 case (“confirmed” exposure; expected to provide a specific indicator, although with low sensitivity since few exposures are notified). Conditional logistic regression models again defined strata by clinic-based or outreach test setting and controlled for all covariates included in primary analyses.

Multivariate-adjusted analyses were subset to individuals who participated in the primary study and thus completed the full telephone-based survey. Expanded data (including pneumococcal sub-study participants) are presented separately, where applicable, for analyses of pneumococcal and SARS-CoV-2 co-detection without covariate adjustment. Missing covariate data where participants in the primary study responded to questions with “Don’t know” or “Refuse” were populated by multiple imputation with five pseudo-datasets. We conducted analyses in R version 4.1.3.

## RESULTS

We enrolled 1,309 participants, including 1,115 in the primary study among farmworkers and 194 in the pneumococcal sub-study only. Saliva specimens were obtained from 1,306 individuals. We excluded 8 samples from individuals who participated in the primary study but did not meet eligibility criteria, and 15 samples due to quality concerns (insufficient volume, label mismatch, or contamination). Additionally, SARS-CoV-2 testing results were inconclusive or unavailable for 5 participants. Final analyses included 1,278 participants (1,095 in the primary study and 183 in the expanded sub-study). Of this sample, 564 and 714 participants were enrolled in clinic-based and mobile outreach testing venues, respectively, among whom 35.7% (199/558, excluding participants missing responses) and 25.4% (181/714), respectively, reported experiencing any solicited clinical symptoms within two weeks before testing (**Table S2**; **Table S3**).

Pneumococcal carriage prevalence was 9.2% (117/1,278) overall (11.2% [63/564] and 7.6% [54/714] in clinic-based and outreach testing, respectively; **Table 1**; **Table S4**). Within the clinic-based sample, prevalence of SARS-CoV-2 infection was 41.3% (26/63) among pneumococcal carriers and 13.8% (69/501) among non-carriers. Within the outreach sample, prevalence of SARS-CoV-2 infection was 11.1% (6/54) among pneumococcal carriers and 6.5% (43/660) among non-carriers. Similar patterns held in analyses defining pneumococcal carriage according to a *lytA* and *piaB* C_*T*_ <35 criterion, with SARS-CoV-2 detected among 34.0% (16/47) of pneumococcal carriers and 10.8% (113/1,048) of non-carriers. Higher point estimates of SARS-CoV-2 infection prevalence among pneumococcal carriers held within strata defined by age, sex, language, country of birth, community of residence, exposure to household crowding, income, education level, occupation, housing type, BMI, cigarette smoking, COVID-19 risk behaviors, and presence or absence of symptoms (**Table 2**).

**Table 1:** Co-detection of SARS-CoV-2 and pneumococcus within the primary and expanded study populations.

| Pneumococcal detection threshold | Population | SARS-CoV-2 infected/Total, *n* (%) |  |  |  |  |  |
| --- | --- | --- | --- | --- | --- | --- | --- |
|  |  | *All testing venues* |  | *Clinic-based testing* |  | *Outreach-based testing* |  |
|  |  | <u>Pneumococcus detected</u> | <u>Pneumococcus not detected</u> | <u>Pneumococcus detected</u> | <u>Pneumococcus not detected</u> | <u>Pneumococcus detected</u> | <u>Pneumococcus not detected</u> |
| *lytA* C<sub>T</sub><40 and *piaB* C<sub>T</sub><40 | Primary study participants | 27/103 (26.2) | 102/992 (10.3) | 25/60 (41.7) | 69/489 (14.1) | 2/43 (4.7) | 33/503 (6.6) |
|  | Expanded dataset, including pneumococcal sub-study participants | 32/117 (27.4) | 112/1,161 (9.6) | 26/63 (41.3) | 69/501 (13.8) | 6/54 (11.1) | 43/660 (6.5) |
| *lytA* C<sub>T</sub><35 and *piaB* C<sub>T</sub><35 | Primary study participants | 16/47 (34.0) | 113/1,048 (10.8) | 15/30 (50.0) | 79/519 (15.2) | 1/17 (5.9) | 34/529 (6.4) |
|  | Expanded dataset, including pneumococcal sub-study participants | 19/53 (35.8) | 125/1,225 (10.2) | 16/31 (51.6) | 79/533 (14.8) | 3/22 (13.6) | 46/692 (6.6) |

SARS-CoV-2: Severe acute respiratory syndrome coronavirus 2, as detected by clinical transcription-mediated amplification testing from oropharyngeal specimens. Pneumococcal detection was defined as C<sub>T</sub> values below 40 for both the *lytA* and *piaB* targets.

**Table 2:**
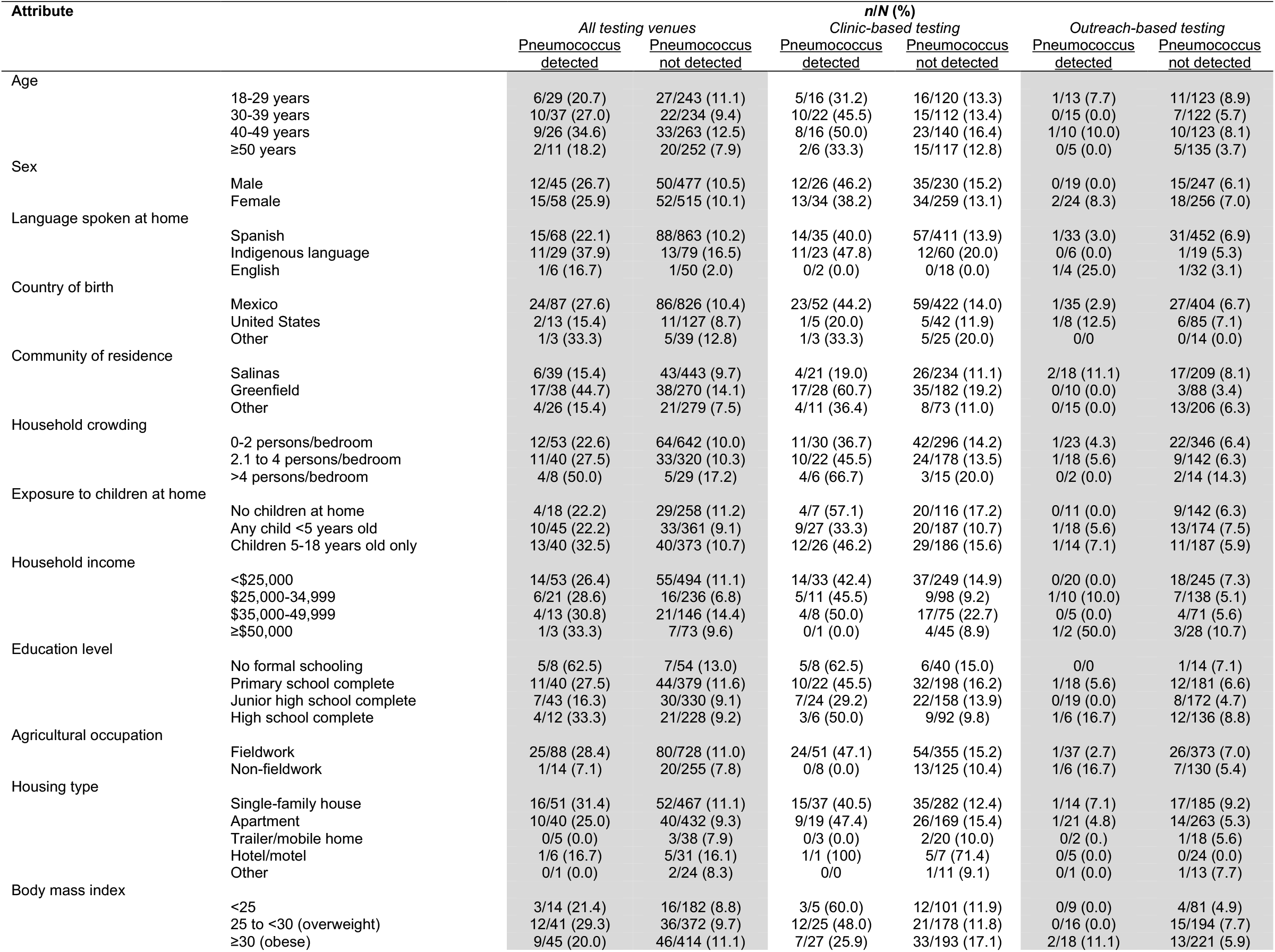
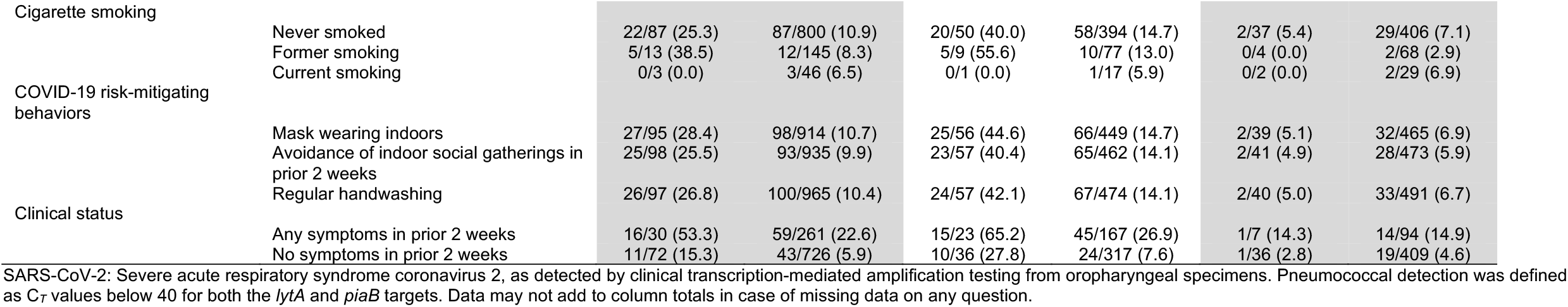
SARS-CoV-2 infection within the primary study population, by pneumococcal carriage status.

Adjusted odds of SARS-CoV-2 infection were 2.73 (1.58-4.69) fold higher among pneumococcal carriers than non-carriers (**Table 3**). Corresponding aOR estimates among participants enrolled in clinic-based and outreach testing were 4.01 (2.08-7.75) and 0.95 (0.20-4.44), respectively. In expanded analyses including participants recruited into the pneumococcal sub-study, the unadjusted OR of SARS-CoV-2 infection associated with pneumococcal carriage in outreach-based test settings was 1.80 (0.73-4.45; **Table S5**), whereas associations within other strata more closely resembled those estimated within the primary study population.

**Table 3:** Association of SARS-CoV-2 infection with pneumococcal carriage detection within the primary study population.

| Participants | All testing venues |  | Clinic-based testing |  | Outreach-based testing |  |
| --- | --- | --- | --- | --- | --- | --- |
|  | Unadjusted OR<br>(95% CI) | Adjusted OR<br>(95% CI) | Unadjusted OR<br>(95% CI) | Adjusted OR<br>(95% CI) | Unadjusted OR<br>(95% CI) | Adjusted OR<br>(95% CI) |
| Total sample | 2.94 (1.79, 4.81) | 2.73 (1.58, 4.69) | 4.33 (2.45, 7.68) | 4.01 (2.08, 7.75) | 0.69 (0.16, 3.06) | 0.95 (0.20, 4.44) |
| Any symptoms in prior two weeks | 3.54 (1.62, 7.79) | 3.38 (1.35, 8.40) | 4.86 (1.95, 12.2) | 6.12 (1.87, 20.5) | — | — |
| No symptoms in prior two weeks | 2.74 (1.35, 5.62) | 2.79 (1.26, 6.24) | 4.61 (2.01, 10.7) | 5.63 (1.91, 16.2) | 0.58 (0.08, 4.53) | 0.78 (0.09, 6.65) |

SARS-CoV-2: Severe acute respiratory syndrome coronavirus 2, as detected by clinical transcription-mediated amplification testing from oropharyngeal specimens; OR: odds ratio; CI: confidence interval. Pneumococcal detection was defined as $C_T$ values below 40 for both the *lytA* and *piaB* targets.

Among participants within the primary study reporting any solicited symptoms, pneumococcal carriage was associated with 3.38 (1.35-8.40) fold higher adjusted odds of SARS-CoV-2 infection, while among participants reporting no symptoms, pneumococcal carriage was associated with 2.79 (1.26-6.24) fold higher adjusted odds of SARS-CoV-2 infection (**Table 3**). The strongest association between pneumococcal carriage and SARS-CoV-2 infection was apparent among symptomatic individuals enrolled in clinic-based testing (aOR=6.12 [1.87-20.5]). However, our analyses did not identify specific clinical symptoms associated with pneumococcal carriage or co-detection with SARS-CoV-2 (**Table S6**; **Table S7**).

Among pneumococcal carriers within the primary study with *piaB* C_*T*_ values of 35-39.9, 30-34.9, and <30, prevalence of SARS-CoV-2 infection was 21.2% (11/52), 26.7% (8/30), and 38.1% (8/21; **Table 4**). Relative to participants who did not carry pneumococcus, adjusted odds of SARS-CoV-2 infection were 1.73 (0.80-3.74), 3.38 (1.35-8.40), and 5.77 (2.06-16.4) fold higher among those who carried pneumococci with *piaB* C_*T*_ values of 35-39.9, 30-34.9, and <30, respectively. Defining *piaB* C_*T*_ values as a continuous exposure, adjusted odds of SARS-CoV-2 infection increased 1.24 (1.00-1.55) fold for each 1-C_*T*_ decrease among participants carrying pneumococci. In agreement with this pattern, the association of SARS-CoV-2 infection with pneumococcal carriage was strengthened when applying a more stringent definition of pneumococcal carriage as *lytA* and *piaB* C_*T*_ <35 (aOR=4.53 [2.20-9.41] vs. 2.73 [1.58-4.69] for *lytA* and *piaB* C_*T*_ <40; **Table S8**). Although C_*T*_ values were positively correlated for *lytA* and *piaB*, we did not identify consistent associations of SARS-CoV-2 infection with *lytA* C_*T*_ values among pneumococcal carriers (**Table 4**; **Figure S1**). Likewise, C_*T*_ values for the SARS-CoV-2 N and E genes in saliva specimens were not associated with pneumococcal detection or with *lytA* and *piaB* C_*T*_ values (**Table S9**; **Figure S2**).

**Table 4:** Association of SARS-CoV-2 infection with *lytA* and *piaB*Cc_*T*_ values in culture-enriched saliva.

| qPCR target | Carriage status and Ct value for indicated qPCR target | SARS-CoV-2 infected/Total, % |  | Odds ratio (95% CI), computed within the primary study population |  |
| --- | --- | --- | --- | --- | --- |
|  |  | Primary study participants | Expanded dataset, including pneumococcal sub-study participants | Unadjusted | Adjusted |
| *lytA* | No pneumococcal carriage | 102/992 (10.3) | 112/1,161 (9.6) | ref. | ref. |
|  | 35-39.9 (with *piaB* C<sub>T</sub> <40) | 5/17 (29.4) | 6/21 (28.6) | 3.66 (1.23, 10.8) | 2.88 (0.86, 9.74) |
|  | 30-34.9 (with *piaB* C<sub>T</sub> <40) | 7/31 (22.6) | 7/32 (21.9) | 2.33 (0.96, 5.63) | 2.26 (0.88, 5.84) |
|  | <30 (with *piaB* C<sub>T</sub> <40) | 15/55 (27.3) | 19/64 (29.7) | 3.11 (1.64, 5.88) | 2.95 (1.47, 5.92) |
|  | Continuous OR—per 1-unit decrease in *lytA* C<sub>T</sub>, among pneumococcal carriers | 27/103 (26.2) | 32/117 (27.4) | 0.97 (0.88, 1.07) | 0.94 (0.81, 1.09) |
| *piaB* | No pneumococcal carriage | 102/992 (10.3) | 112/1,161 (9.6) | ref. | ref. |
|  | 35-39.9 (with *lytA* C<sub>T</sub> <40) | 11/52 (21.2) | 13/58 (22.4) | 2.24 (1.10, 4.58) | 1.73 (0.80, 3.74) |
|  | 30-34.9 (with *lytA* C<sub>T</sub> <40) | 8/30 (26.7) | 9/33 (27.3) | 2.94 (1.26, 6.90) | 3.38 (1.35, 8.40) |
|  | <30 (with *lytA* C<sub>T</sub> <40) | 8/21 (38.1) | 10/26 (38.5) | 5.03 (1.98, 12.8) | 5.77 (2.06, 16.4) |
|  | Continuous OR—per 1-unit decrease in *piaB* C<sub>T</sub>, among pneumococcal carriers | 27/103 (26.2) | 32/117 (27.4) | 1.07 (0.96, 1.19) | 1.24 (1.00, 1.55) |

SARS-CoV-2: Severe acute respiratory syndrome coronavirus 2, as detected by clinical transcription-mediated amplification testing from oropharyngeal specimens; OR: odds ratio; CI: confidence interval. Pneumococcal detection was defined as C<sub>T</sub> values below 40 for both the *lytA* and *piaB* targets.

Among participants in the primary study who reported no suspected or notified SARS-CoV-2 exposure within 2 weeks before testing, prevalence of SARS-CoV-2 infection was 19.2% (15/78) among pneumococcal carriers and 7.9% (61/769) among non-carriers (**Table 5**). In contrast, 69.2% (9/13) of pneumococcal carriers and 26.1% (30/115) of non-carriers who reported suspected exposure tested positive for SARS-CoV-2 infection.

**Table 5:** Association of SARS-CoV-2 infection with recent exposure among individuals carrying and those not carrying pneumococcus.

| Exposure status | SARS-CoV-2 infected/Total (%), by pneumococcal carriage status |  | Pneumococcal carriage not detected |  | Pneumococcal carriage detected |  |
| --- | --- | --- | --- | --- | --- | --- |
|  | Pneumococcus detected | Pneumococcus not detected | Unadjusted OR (95% CI) | Adjusted OR (95% CI) | Unadjusted OR (95% CI) | Adjusted OR (95% CI) |
| No contact with symptomatic individual in prior two weeks | 15/78 (19.2) | 61/769 (7.9) | ref. | ref. | ref. | ref. |
| Contact with symptomatic individual in prior two weeks—Any | 9/13 (69.2) | 30/115 (26.1) | 3.21 (1.95, 5.26) | 3.29 (1.94, 5.59) | 7.25 (1.95, 26.8) | 7.64 (1.91, 30.7) |
| Contact with symptomatic individual in prior two weeks—Home | 6/9 (66.7) | 18/61 (29.5) | 3.82 (2.10, 6.99) | 3.99 (2.06, 7.67) | 5.35 (1.22, 24.0) | 6.66 (1.32, 33.0) |
| Contact with symptomatic individual in prior two weeks—Work | 5/7 (71.4) | 16/63 (25.4) | 2.49 (1.33, 4.63) | 2.26 (1.17, 4.38) | 6.34 (1.14, 34.8) | 6.25 (1.00, 38.0) |
| No notification of contact with confirmed case in prior two weeks | 25/98 (25.5) | 94/922 (10.2) | ref. | ref. | ref. | ref. |
| Notification of contact with confirmed case in prior two weeks | 2/5 (40.0) | 8/70 (11.4) | 0.90 (0.42, 1.98) | 0.83 (0.37, 1.85) | 2.51 (0.37, 17.1) | 2.14 (0.28, 16.0) |

SARS-CoV-2: Severe acute respiratory syndrome coronavirus 2, as detected by clinical transcription-mediated amplification testing from oropharyngeal specimens; OR: odds ratio; CI: confidence interval. Pneumococcal detection was defined as $C_T$ values below 40 for both the *lytA* and *piaB* targets.

The aOR of SARS-CoV-2 infection associated with suspected exposure were 7.64 (1.91-30.7) among pneumococcal carriers and 3.29 (1.94-5.59) among non-carriers. These differences in aOR point estimates held within analyses restricted to participants reporting such exposures in the home (aOR=6.66 [1.32-33.0] for carriers and 3.99 [2.06-7.67] for non-carriers) and workplace (aOR=6.25 [1.00-38.0] for carriers and 2.26 [1.17-4.38] for non-carriers). A similar pattern was apparent for participants who reported that they were notified of exposure to a confirmed COVID-19 case, although such analyses were underpowered due to the low frequency of notified exposures. The aOR of SARS-CoV-2 infection associated with notified exposure was 2.14 (0.28-16.0) among pneumococcal carriers and 0.83 (0.37-1.85) among non-carriers.

## DISCUSSION

We identify a synergistic association between pneumococcal carriage and SARS-CoV-2 infection among adults tested in ambulatory clinics and community settings. This association was strongest among participants recruited through clinical SARS-CoV-2 testing and those experiencing acute COVID-19 symptoms. Among pneumococcal carriers, adjusted odds of concurrent SARS-CoV-2 infection increased 24% for each 1-unit decrease in *piaB* C_*T*_, indicating a density-dependent relationship. Last, the increase in odds of SARS-CoV-2 infection associated with recent exposure to a symptomatic individual was roughly two-fold higher among pneumococcal carriers as compared to non-carriers. Collectively, our findings are consistent with the hypothesis of interactions between pneumococci and SARS-CoV-2 in the upper respiratory tract.

In agreement with our findings, a prior study reported that pneumococcal carriage was detected more often among adults with asymptomatic or mild SARS-CoV-2 infection than among uninfected adults, and was associated with reduced cellular and mucosal immune responses to SARS-CoV-2.^19^ Similar interactions have been reported in studies involving other respiratory viruses,^4–11^ which have further documented increased pneumococcal density in the upper airway during viral infection.^5,7–9^ While cross-sectional epidemiologic studies cannot distinguish interaction mechanisms underlying these findings—such as pneumococcal outgrowth during viral infection, or facilitation of viral infection during pneumococcal carriage—several studies in mouse models have suggested facilitative interactions may be bidirectional. Challenge with wild-type influenza A virus or live attenuated influenza vaccine extended the duration of pneumococcal carriage in mice, regardless of whether exposure to pneumococci preceded or followed exposure to influenza.^12^ Additionally, pneumococcal/SARS-CoV-2 coinfection was associated with enhanced mortality in mice, regardless of the order of challenge with SARS-CoV-2 and pneumococci.^31^ Last, in humans, experimentally-induced pneumococcal colonization dampened mucosal responses to subsequent challenge with live attenuated influenza vaccine.^13^

In contrast to the observed associations of pneumococcal density with SARS-CoV-2 presence or absence, SARS-CoV-2 C_*T*_ values were not associated with pneumococcal carriage or density in our study. This circumstance may owe to pronounced temporal variability in SARS-CoV-2 viral load over the course of infection.^32^ Importantly, our study could not account for time since SARS-CoV-2 acquisition, which is a primary determinant of viral load and may have influenced the findings.

Associations of recent suspected or notified SARS-CoV-2 exposure with risk of infection were stronger among pneumococcal carriers than non-carriers in our study. As typical durations of pneumococcal carriage among adults exceed the 2-week recall period of this exposure,^33^ pneumococcal colonization likely preceded SARS-CoV-2 exposure for most study participants with both pathogens detected. At least two interpretations of this finding are possible. First, our finding may signify heightened risk of SARS-CoV-2 infection, given exposure, in the context of nasopharyngeal inflammation induced by pneumococcal colonization. In support of this hypothesis, pre-existing pneumococcal carriage has been found to predict increased risk of new-onset of virus-associated acute respiratory infections among children.^15^ In another study among children, pneumococcal carriage density was found to increase markedly with acquisition of RSV or rhinoviruses,^34^ although such increases could also be driven by a loss of control of existing pneumococcal colonization following viral acquisition.^35^ An alternative interpretation may thus view our finding as an artifact of enhanced pneumococcal outgrowth associated with recent SARS-CoV-2 exposure and acquisition. If such outgrowth is transient in nature, higher pneumococcal density (and thus pneumococcal carriage detection) could be associated with more recent SARS-CoV-2 acquisition, thereby accounting for stronger associations between SARS-CoV-2 infection and exposure in the prior 2 weeks among individuals found to carry pneumococci. As our cross-sectional study design cannot distinguish between these hypotheses, longitudinal studies relating baseline pneumococcal carriage status to risk of subsequent viral acquisition are needed.

Our study has several limitations. Recruitment of farmworkers in low-income communities indicates that absolute estimates of prevalence of pneumococcal carriage and SARS-CoV-2 infection are not externally generalizable. Our data do not encompass potential social linkages among participants, who in some instances may have lived or worked together. As recruitment for this study occurred in summer and fall of 2020, cases in our study were not infected with recent SARS-CoV-2 variants of concern. While our analysis demonstrated density-dependent relationships between pneumococcal carriage and SARS-CoV-2 infection based on *piaB* C_*T*_ values, these readings were taken following a culture enrichment step. Measures of bacterial abundance obtained prior to culture enrichment would be expected to better represent carriage density in the host, although previous studies have demonstrated strong correlations in pneumococcal C_*T*_ values before and after culture enrichment.^36,37^ Last, our study population was recruited in ambulatory care settings and outreach testing venues, and does not represent the full spectrum of COVID-19 disease severity including cases requiring hospital or intensive care unit admission. Observed associations should thus be interpreted as markers of pathogen interactions either in mild-to-moderate infections or at early stages in the clinical course, prior to severe disease progression.

Our findings corroborate several previous lines of evidence^19–21,31^ suggesting a relationship exists between pneumococci and SARS-CoV-2 infection among adults. Whereas IPD incidence declined precipitously during the early COVID-19 pandemic, this pattern may have been driven, in part, by frequent antibiotic treatment of COVID-19 patients who would otherwise be at risk,^17^ absent transmission of other viruses. Of note, non-bacteremic pneumococcal pneumonia and LRTI have persisted during the pandemic, and pneumococcal antigen detection has been associated with increased disease severity among hospitalized COVID-19 cases.^18^ Increased density may facilitate pneumococcal transmission during SARS-CoV-2 infection in addition to increasing individuals’ risk of subsequent LRTI and IPD, absent antibiotic treatment.^38,39^ Monitoring for severe co-infections involving SARS-CoV-2 and pneumococcus thus remains warranted as antimicrobial stewardship for COVID-19 cases improves. While vaccines now offer a primary prevention strategy for COVID-19, our results pinpoint interactions involving pneumococci in the upper airway as a component of SARS-CoV-2 natural history, resembling observations with other viruses. These findings may inform control strategies against polymicrobial infections.

## Data Availability

Produced in the present study are available upon reasonable request to the authors, for investigators who will collaborate with the investigators in secondary analyses under the oversight of applicable UC Berkeley ethics committees.

## Text S1: Supplemental laboratory methods

### Culture enrichment and bacterial DNA extraction

Raw saliva specimens were frozen at –20ºC after collection and thereafter stored at –80ºC. Samples were thawed on ice after arrival at the laboratory; after thawing, 100μL of unprocessed saliva was spread onto trypticase soy agar supplemented with 7% sheep’s blood and 5mg/L gentamicin. Plates were then incubated at 37ºC with 5% CO_2_ for 14-19 hours. Bacterial growth was harvested into 2.1mL of 10% glycerol brain-heart infusion broth (BHI), and stored at –20ºC. Specimen DNA was extracted from 200μL of the culture-enriched sample after being thawed, vortexed, and transferred to sterile 1.5mL Eppendorf tubes prefilled with freshly prepared lysis buffer (40mg/mL lysozyme and 75U/mol mutanolysin) for 90 minutes’ incubation at 37ºC. Next, 20μL of protease K was added to each sample, followed by incubation for 60 minutes at 56ºC. Subsequent extraction steps used the QIAGEN DNeasy Blood and Tissue Kit (QIAGEN; Hilden, Germany) following manufacturer’s instructions. Briefly, DNA was eluted into 50μL elution buffer and stored at –20ºC; samples with DNA concentration <20ng/μL, measured using a spectrophotometer, underwent a second extraction. Positive and negative control DNA was extracted from pure cultures of *S. pneumoniae* by the boilate method. Briefly, colonies grown overnight on a TSA-GENT plate were extracted and suspended in 60μL nuclease-free water, then placed into a water bath heated to 100ºC. Samples were boiled for 10 minutes before being centrifuged for 10 minutes at 14,000 rpm. Nucleic acid concentration was measured using a spectrophotometer following removal of the supernatant. Control DNA was diluted with nuclease-free water to a concentration of 50ng/μL.

### Detection of lytA and piaB in culture-enriched specimens

Positive control samples used a diluted series of the reference strain ATCC 49619, while negative controls were DNA from human saliva previously characterized as negative for *S. pneumoniae*, and DNA from *E. coli* reference strain ATCC 25922. Gene detection via qPCR was conducted with primer concentrations at 400nM and 300nM, and probe concentrations at 75nM and 200nM, for *lytA* and *piaB*, respectively. All qPCR assays tested 2.5μL of DNA template in 25μL reactions. Thermal cycling conditions were as follows: 3 minutes at 95ºC for initial denaturation; 15 seconds at 98ºC for denaturation; and 30 seconds at 60ºC for annealing.

### Detection of SARS-CoV-2 in saliva samples

Prior to measuring C_*T*_ values, all samples were subjected to a freeze-thaw cycle for aliquoting. In addition, saliva samples from participants with a positive diagnostic test result were heat inactivated at 56ºC for 30 minutes for safety purposes. Saliva samples were defrosted on ice for approximately 30 minutes. Once defrosted, raw saliva was pipetted into Omnigene tubes (Omnigene OM-505) and an equal volume of Omnigene solution was added. Samples were then extracted as previously described.^1^ Briefly, samples were incubated at 50ºC for 2 hours, followed by 30 minutes’ incubation at 65ºC. Using a Hamilton Microlab STARlet, 112μL of processed saliva was combined with 338μL of 2x DNA/RNA Shield (Zymo; R1200). RNA was extracted with the MagMax Viral Pathogen Kit (ThermoFisher; A42352) using a Hamilton Vantage. Viral N and E genes, and human RNaseP were amplified using the LuNER assay, as previously described.^2^ Positive and negative controls were included for extraction and qPCR and were used to establish plate validity. RNaseP amplification within each sample was used to determine sample validity. Samples were re-extracted and re-amplified as needed for final C_*T*_ values.

---

**Table S1:**
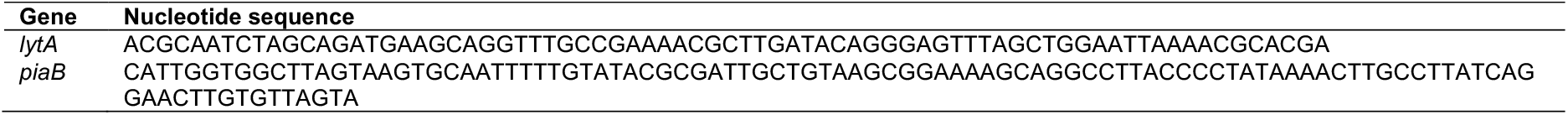
Gene targets for molecular detection of pneumococci.

**Table S2:**
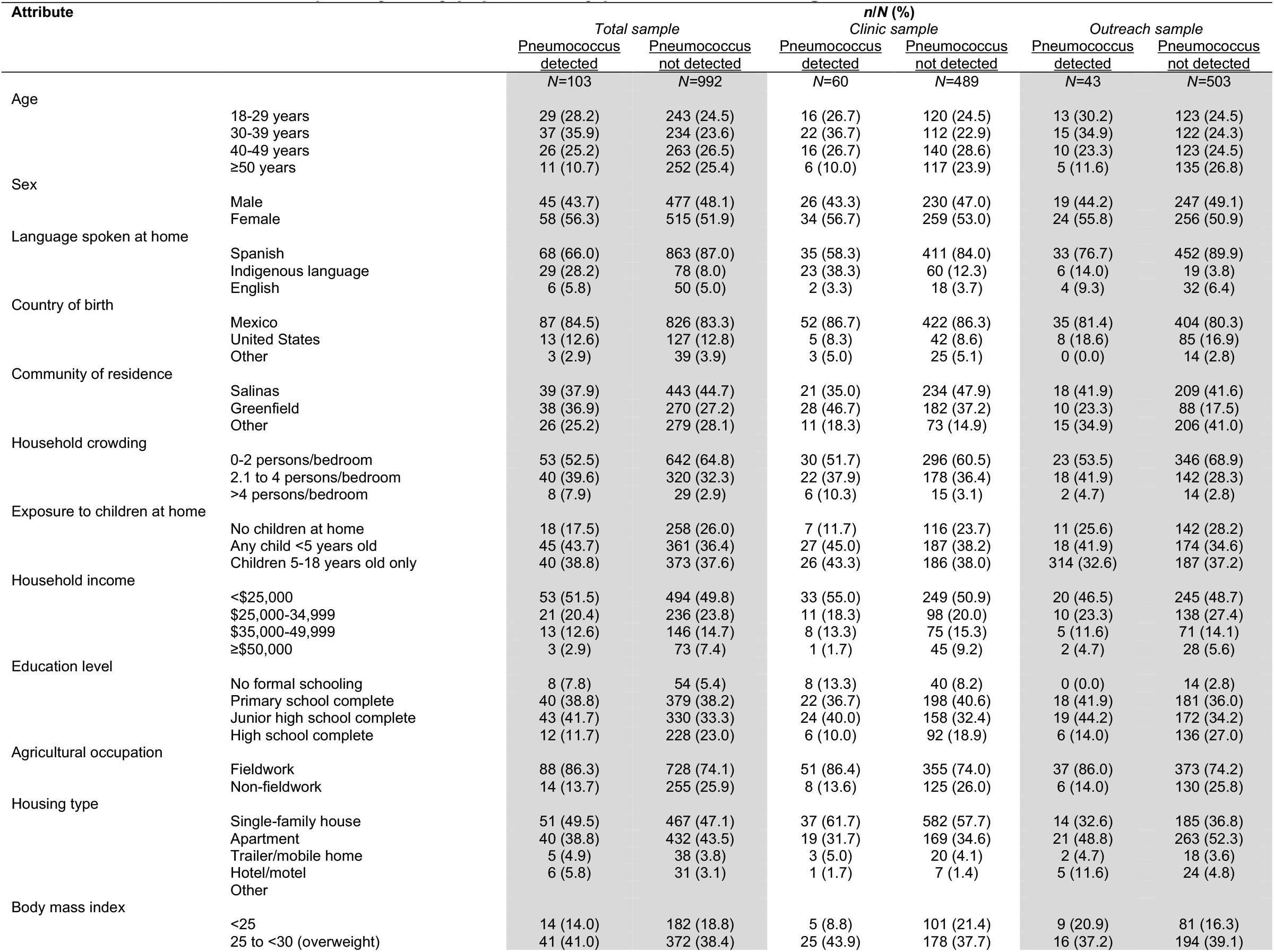
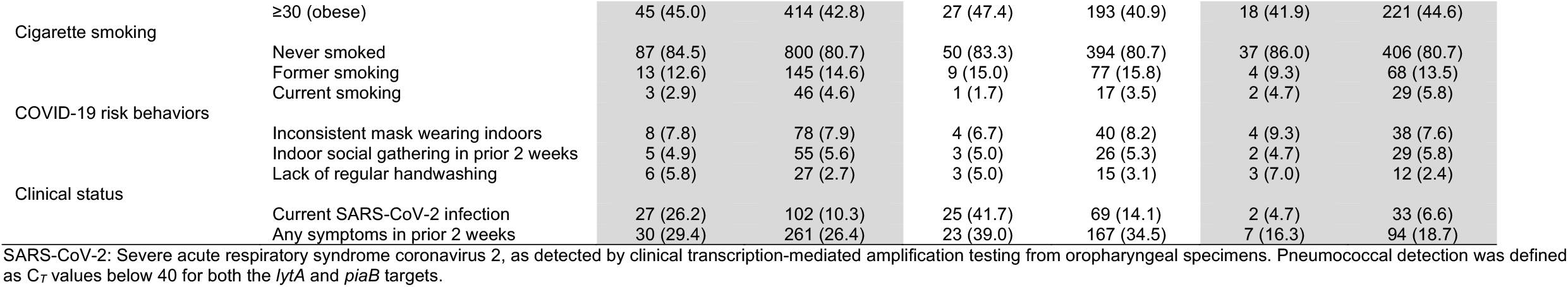
Characteristics of the primary study population, by pneumococcal carriage status.

**Table S3:**
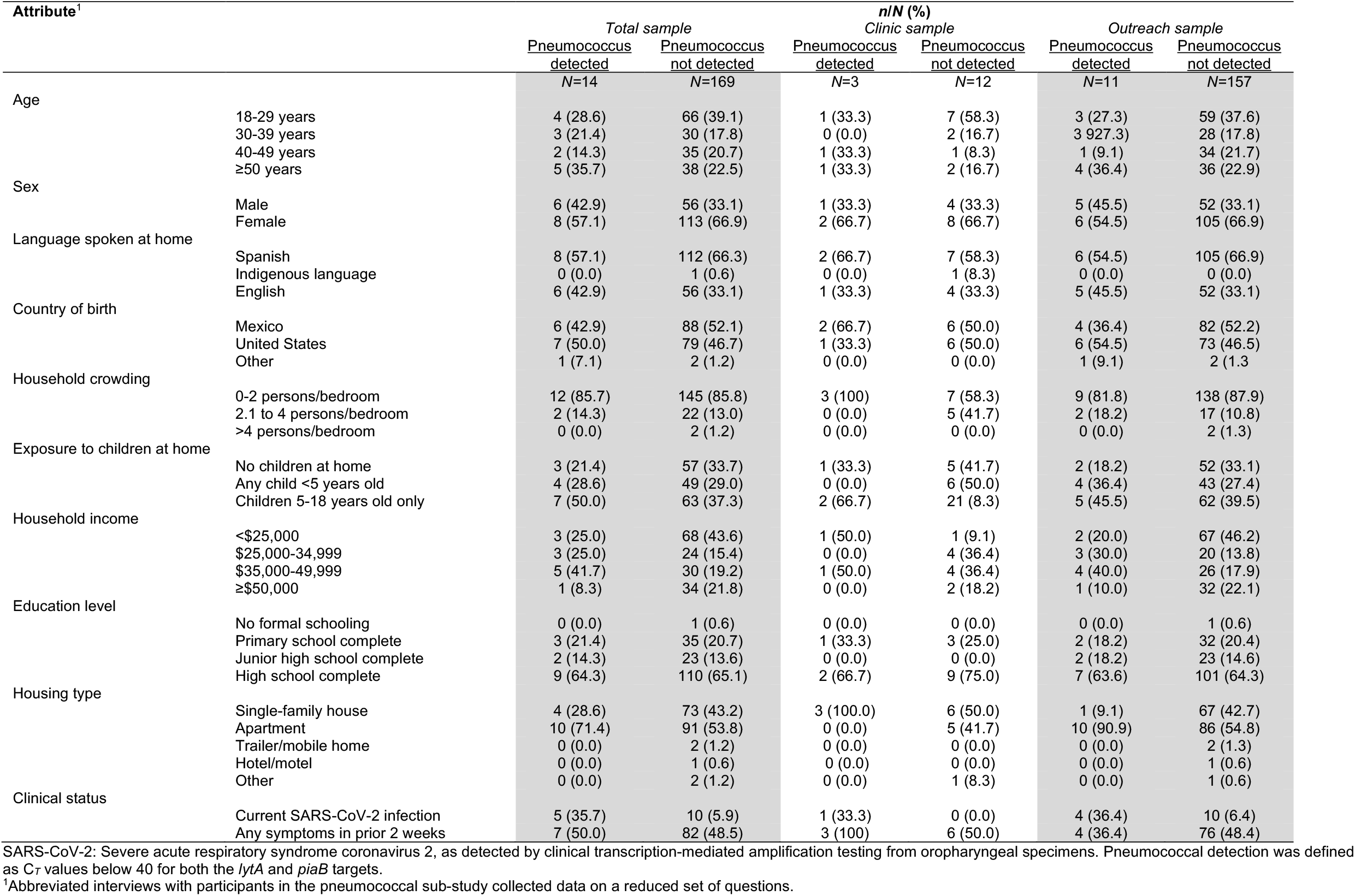
Characteristics of the pneumococcal sub-study population, by pneumococcal carriage status.

**Table S4:** Association of SARS-CoV-2 infection with *lytA* detection in *piaB*-negative specimens.

| Sample | Participants | SARS-CoV-2 infected/Total (%) |  |  |  |
| --- | --- | --- | --- | --- | --- |
|  |  | *lytA* detected | *lytA* not detected | Unadjusted OR (95% CI) | Adjusted OR (95% CI)<sup>1</sup> |
| Total sample | Primary study participants | 28/193 (14.5) | 74/793 (9.3) | 1.63 (1.02, 2.61) | 1.57 (0.96, 2.57) |
|  | Expanded dataset, including pneumococcal sub-study participants | 29/218 (13.3) | 83/937 (8.9) | 1.55 (0.98, 2.44) | -- |
| Clinic-based sample | Primary study participants | 23/99 (23.2) | 46/387 (11.9) | 2.24 (1.29, 3.91) | 1.95 (1.07, 3.56) |
|  | Expanded dataset, including pneumococcal sub-study participants | 23/100 (23.0) | 46/398 (11.6) | 2.28 (1.30, 3.98) | -- |
| Outreach-based sample | Primary study participants | 5/94 (5.3) | 28/406 (6.9) | 0.76 (0.29, 2.02) | 0.77 (0.27, 2.18) |
|  | Expanded dataset, including pneumococcal sub-study participants | 6/118 (5.1) | 37/539 (6.9) | 0.73 (0.30, 1.76) | -- |

SARS nCoV-2: Severe acute respiratory syndrome coronavirus 2, as detected by clinical transcription-mediated amplification testing from oropharyngeal specimens; OR: odds ratio. Data are subset to specimens without *piaB* detection. We indicate dashes where models lacked sufficient sample to converge for all parameter estimates.
<sup>1</sup>Adjusted odds ratios were not computed for analyses within the expanded dataset, as data were collected for a limited set of covariates among pneumococcal sub-study participants.

**Table S5:** Unadjusted associations of SARS-CoV-2 infection with pneumococcal carriage detection within the expanded study population, including participants in the primary study and pneumococcal sub-study.

| Participants | All testing venues | Clinic-based testing | Outreach-based testing |
| --- | --- | --- | --- |
|  | Unadjusted OR (95% CI) | Unadjusted OR (95% CI) | Unadjusted OR (95% CI) |
| Total sample | 3.31 (2.08, 5.24) | 4.40 (2.50, 7.72) | 1.80 (0.73, 4.45) |
| Any symptoms in prior two weeks | 4.15 (2.04, 8.42) | 4.40 (1.87, 10.3) | 3.64 (0.99, 13.4) |
| No symptoms in prior two weeks | 3.01 (1.50, 6.01) | 4.73 (2.04, 10.9) | 1.16 (0.26, 5.13) |

SARS-CoV-2: Severe acute respiratory syndrome coronavirus 2, as detected by clinical transcription-mediated amplification testing from oropharyngeal specimens; OR: odds ratio. Pneumococcal detection was defined as $C_T$ values below 40 for both the *lytA* and *piaB* targets.

**Table S6:**
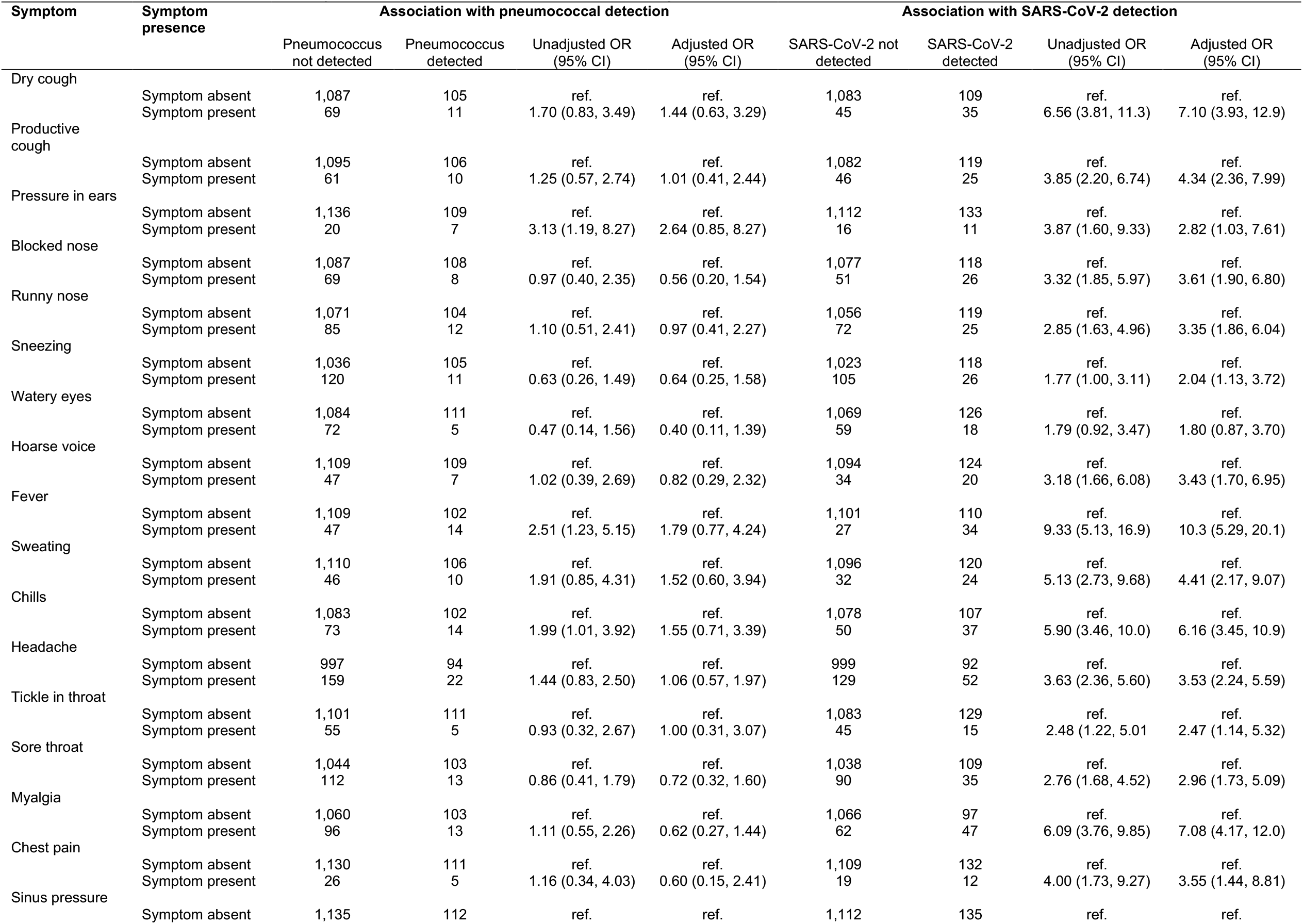
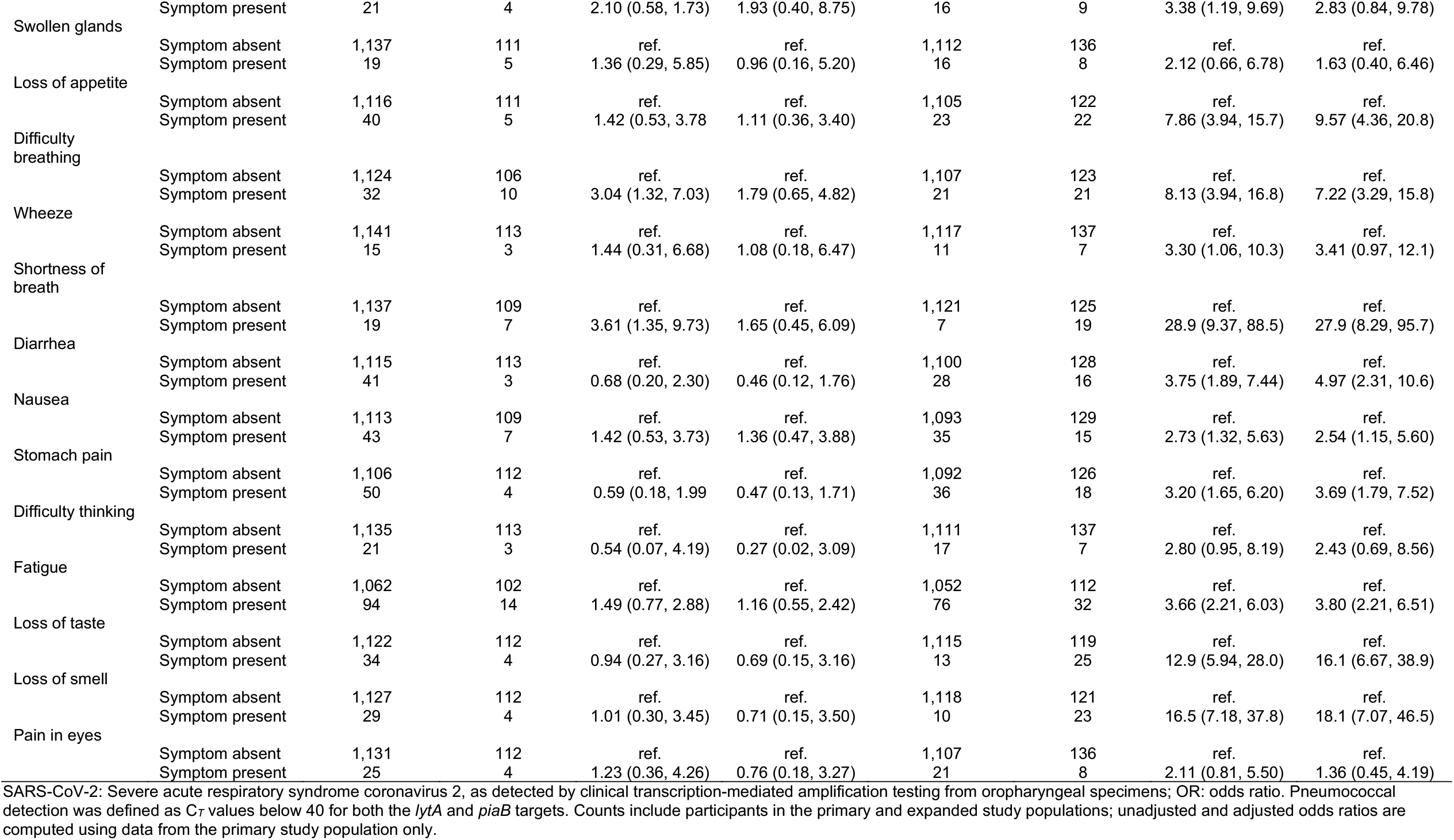
Association of clinical symptoms with pneumococcal carriage detection and SARS-CoV-2 infection.

**Table S7:**
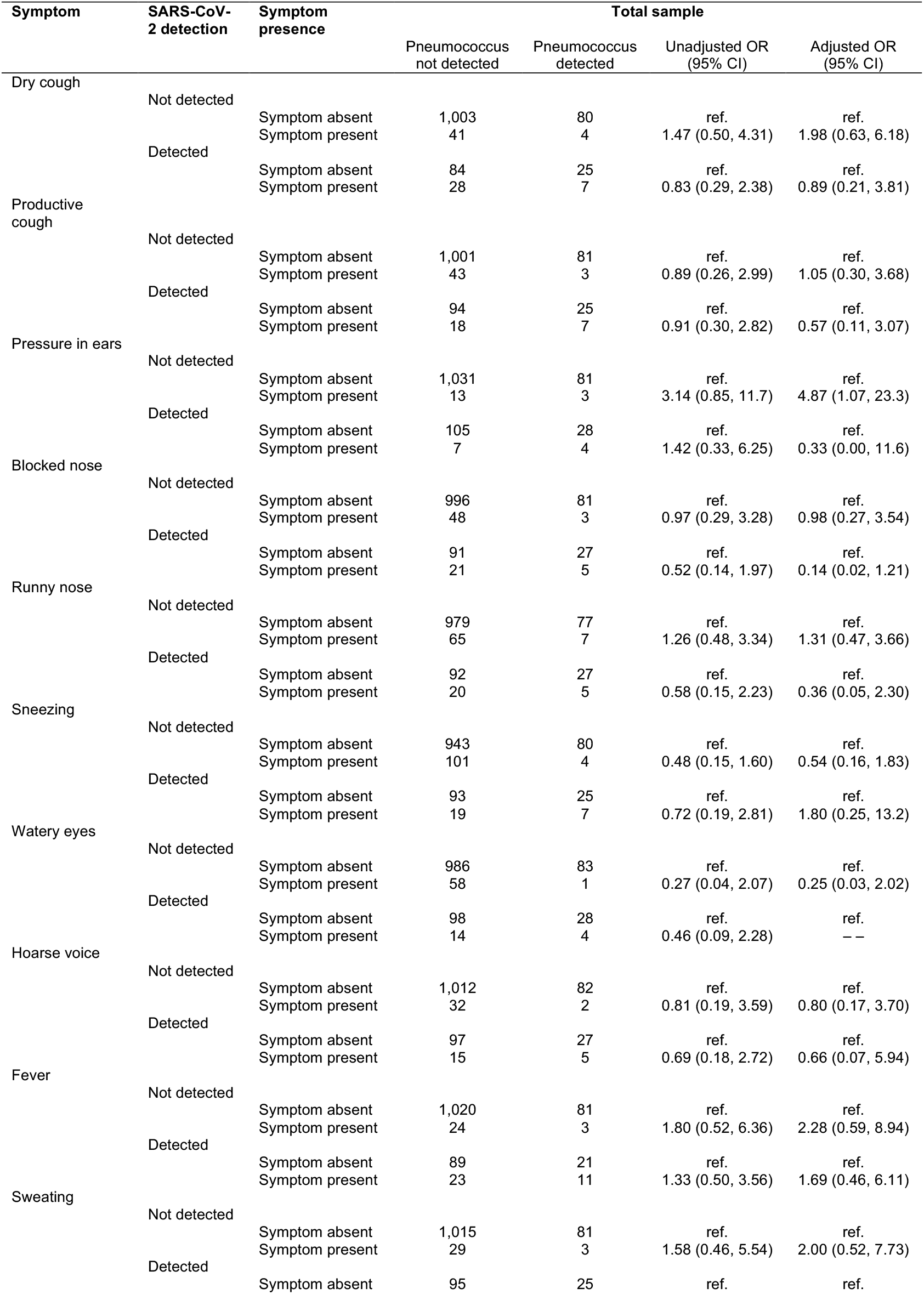
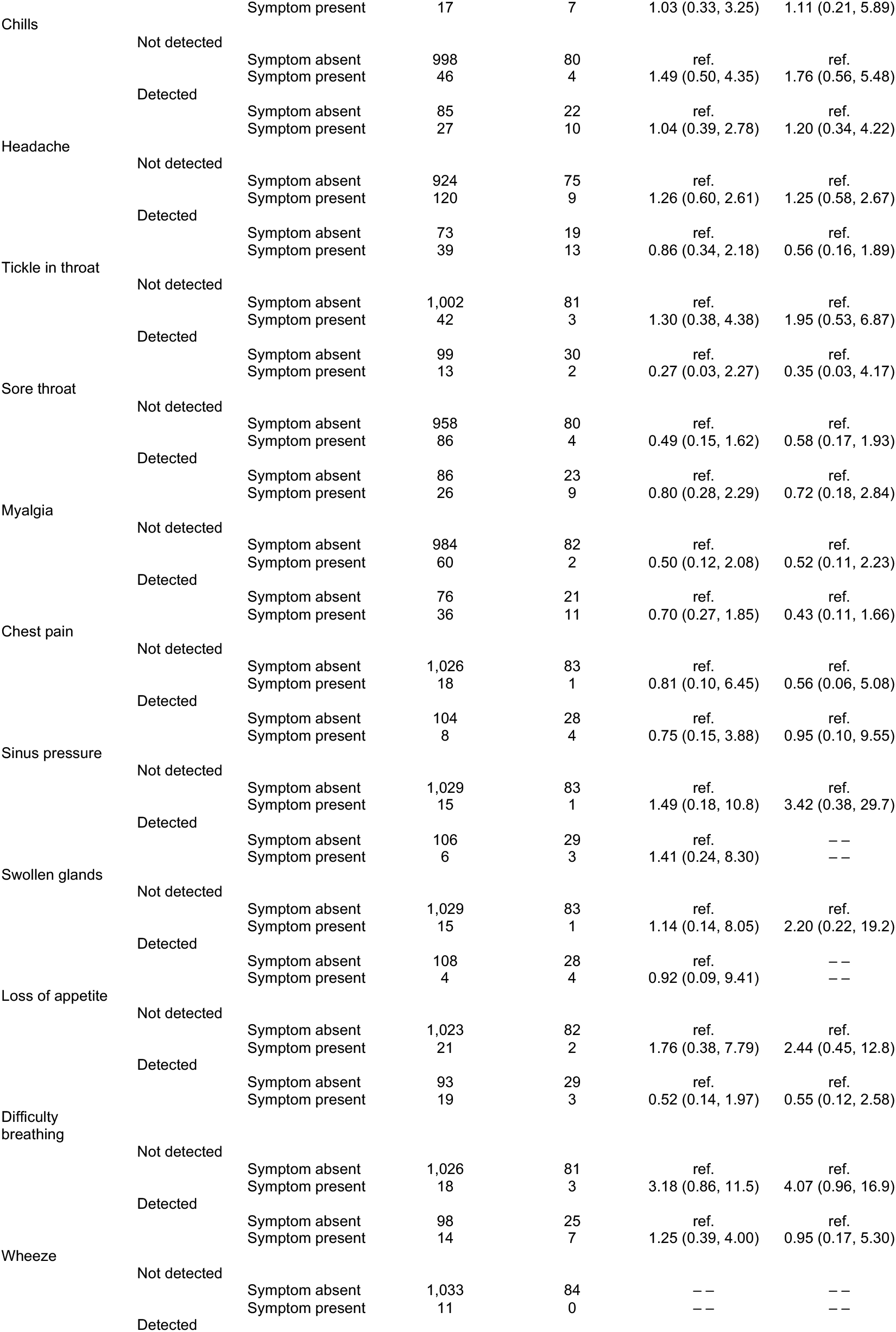
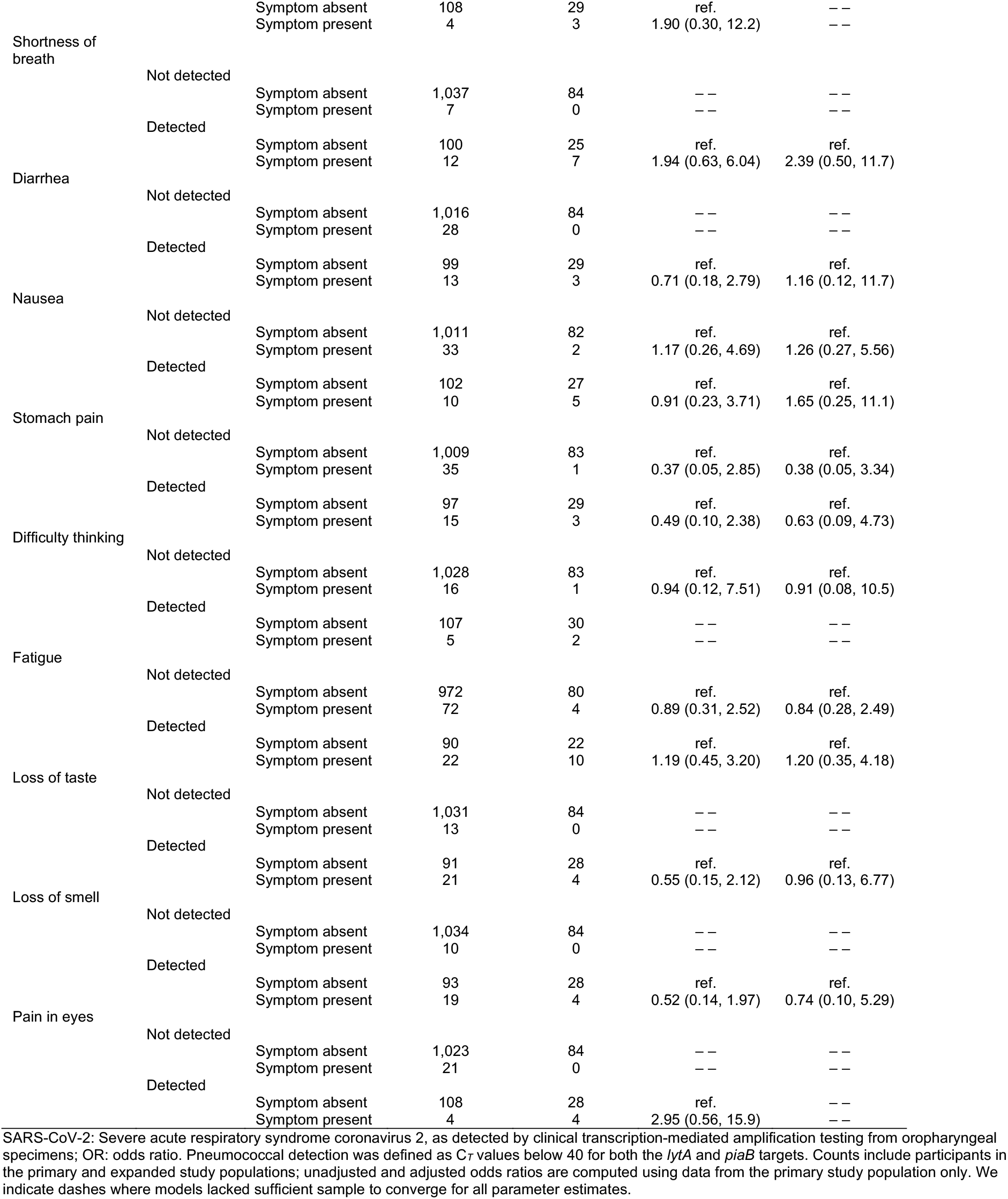
Association of clinical symptoms with detection of pneumococcus, stratifying by SARS-CoV-2 infection status.

**Table S8:**
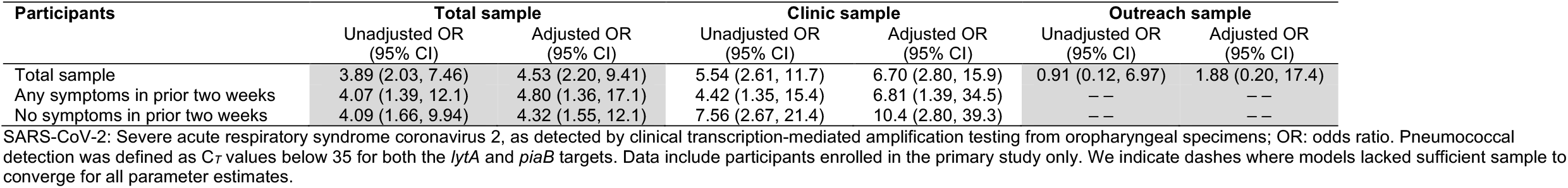
Association of SARS-CoV-2 infection with pneumococcal carriage at *lytA* C_*T*_<35 and *piaB* C_*T*_<35 detection thresholds.

| Participants | Total sample |  | Clinic sample |  | Outreach sample |  |
| --- | --- | --- | --- | --- | --- | --- |
|  | Unadjusted OR<br>(95% CI) | Adjusted OR<br>(95% CI) | Unadjusted OR<br>(95% CI) | Adjusted OR<br>(95% CI) | Unadjusted OR<br>(95% CI) | Adjusted OR<br>(95% CI) |
| Total sample | 3.89 (2.03, 7.46) | 4.53 (2.20, 9.41) | 5.54 (2.61, 11.7) | 6.70 (2.80, 15.9) | 0.91 (0.12, 6.97) | 1.88 (0.20, 17.4) |
| Any symptoms in prior two weeks | 4.07 (1.39, 12.1) | 4.80 (1.36, 17.1) | 4.42 (1.35, 15.4) | 6.81 (1.39, 34.5) | -- | -- |
| No symptoms in prior two weeks | 4.09 (1.66, 9.94) | 4.32 (1.55, 12.1) | 7.56 (2.67, 21.4) | 10.4 (2.80, 39.3) | -- | -- |

SARS-CoV-2: Severe acute respiratory syndrome coronavirus 2, as detected by clinical transcription-mediated amplification testing from oropharyngeal specimens; OR: odds ratio. Pneumococcal detection was defined as C<sub>T</sub> values below 35 for both the *lytA* and *piaB* targets. Data include participants enrolled in the primary study only. We indicate dashes where models lacked sufficient sample to converge for all parameter estimates.

**Table S9:**
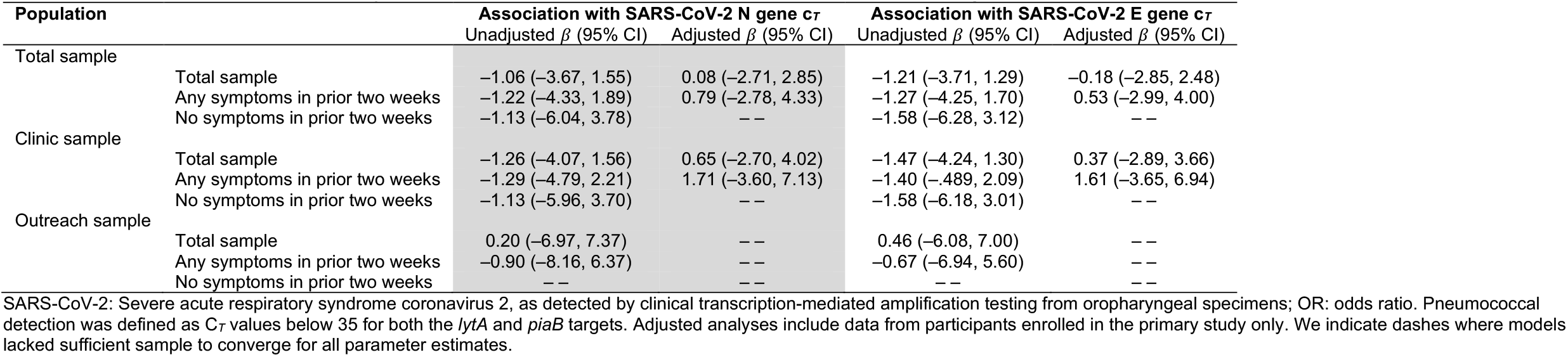
Association of salivary SARS-CoV-2 C_*T*_ values with pneumococcal carriage detection.

**Figure S1:**
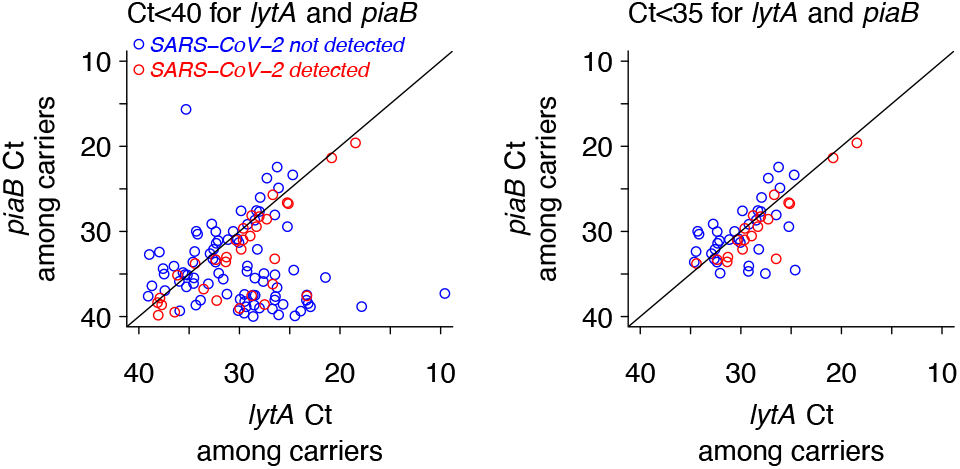
Correlation between *lytA* and *piaB* C*T* values among pneumococcal carriers. Panels illustrate C*T* values for all individuals found to carry pneumococci (based on C*T* for both *lytA* and *piaB*; left) and for individuals with C*T* values below 35 for both *lytA* and *piaB* (right).

**Figure S2:**
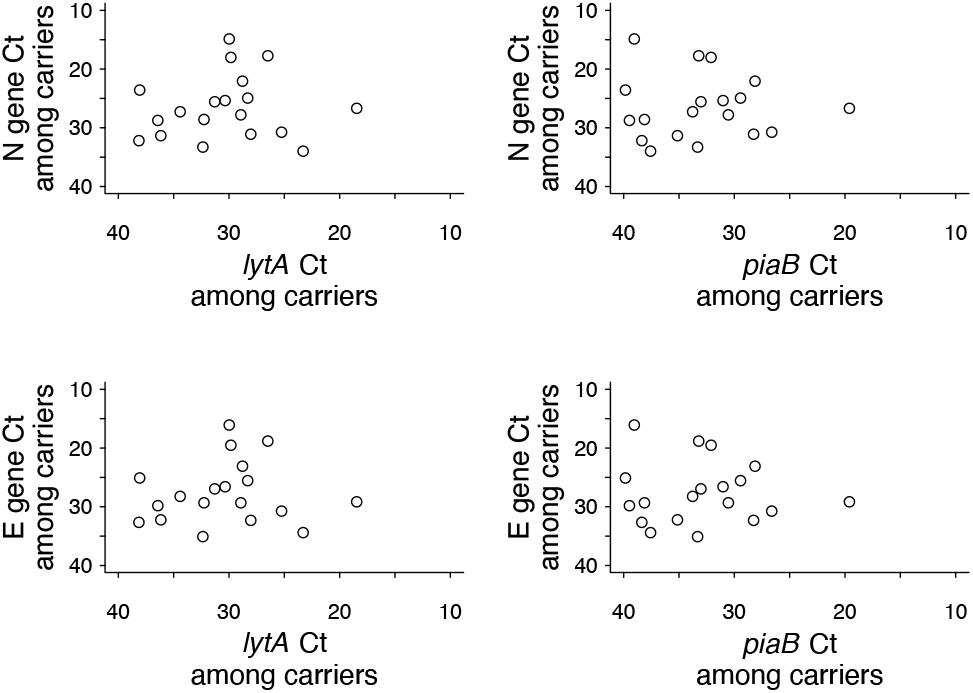
Correlation of qPCR C*T* value for SARS-CoV-2 and pneumococcal gene targets in saliva samples. Panels illustrate C*T* values for the SARS-CoV-2 nucleocapsid (N; top panels) and structural envelope protein (E; bottom panels) genes against c*T* values for the pneumococcal *lytA* (left panels) and *piaB* (right panels) genes. Data are subset to individuals found to be infected with SARS-CoV-2 and to carry pneumococci.

